# Dental mitigation strategies to reduce aerosolization of SARS-CoV-2

**DOI:** 10.1101/2021.03.24.21254254

**Authors:** Jon J. Vernon, Emma V. I. Black, Thomas Dennis, Deirdre A. Devine, Louise Fletcher, David J Wood, Brian R Nattress

## Abstract

Limiting infection transmission is central to the safety of all in dentistry, particularly during the current SARS-CoV-2 pandemic. Aerosol-generating procedures (AGPs) are crucial to the practise of dentistry; it is imperative to understand the inherent risks of viral dispersion associated with AGPs and the efficacy of available mitigation strategies.

In a dental surgery setting, crown preparation and root canal access procedures were performed with an air turbine or electric speed-controlled hand-piece, with mitigation via rubber dam or high-volume aspiration and a no mitigation control. A phantom head was used with a 1.5 mL flow of artificial saliva infected with Φ6 bacteriophage (a surrogate virus for SARS-CoV-2) at ∼10^8^ plaque forming units mL^-1^, reflecting the upper limits of reported salivary SARS-CoV-2 levels. Bioaerosol dispersal was measured using agar settle plates lawned with the bacteriophage’s host, *Pseudomonas syringae*. Viral air concentrations were assessed using MicroBio MB2 air sampling, and particle quantities using Kanomax 3889 GEOα particle counters.

Compared to an air turbine, the electric hand-piece reduced settled bioaerosols by 99.72%, 100.00% and 100.00% for no mitigation, aspiration and rubber dam, respectively. Bacteriophage concentrations in the air were reduced by 99.98%, 100.00% and 100.00%, with the same mitigation strategies. Use of the electric hand-piece with high-volume aspiration, resulted in no detectable bacteriophage, both on settle plates and in air samples taken 6-10-minutes post-procedure.

To our knowledge, this study is the first to report the aerosolization of active virus as a marker for risk determination in the dental setting. Whilst this model represents a worst-case scenario for possible SARS-CoV-2 dispersal, these data showed that the use of electric hand-pieces can vastly reduce the risk of viral aerosolization, and therefore remove the need for clinic fallow time. Furthermore, our findings indicate that the use of particle analysis alone cannot provide sufficient insight to understand bioaerosol infection risk.

## Introduction

The potential nosocomial spread of SARS-CoV-2 coronavirus and other pathogens through oral fluid aerosolization provides a significant risk to the safety of patients, dentists and oral healthcare teams. During the current SARS-CoV-2 pandemic, extensive constraints have been placed on dentistry across the world, with a particular focus on aerosol-generating procedures (AGPs). [1] These constraints impact widely upon how dentistry can be delivered in both dental practices or offices and on multi-occupancy teaching clinics. There is a paucity of robust data supporting some of these restrictions and further research is essential to investigate the efficacy of mitigation strategies and the requirement for fallow time between patients.

In dental settings, aerosols are generated from the use of dynamic dental instruments such as ultrasonic scaling tips, high-speed dental hand-pieces and 3:1 syringes. The aerosols are known to contain saliva/blood, microorganisms (including viruses) from the oral cavity of the patient and are created by air mixture from the hand-piece and water flowing from the dental unit water line. [2-4] This generation of aerosols is considered to be unavoidable due to the use of rapidly rotating high-speed dental hand-pieces. Exposure to aerosol particles of 50µm or smaller pose the greatest risk because the smaller the particle size, the more likely they are to remain airborne for long periods. Therefore, they are more capable of entering the nasal passages and penetrating deep into the respiratory system. [5]

Aerosol composition also varies from patient to patient, the site in which the dental procedures are carried out and the type of procedure performed. Ninety percent of the aerosols generated are known to have a mass median aerodynamic diameter of <5µm, and as the dental environment is known to be contaminated with various microorganisms, the resulting aerosols pose a threat to dental workers. [4, 6] SARS-CoV-2 represents the most significant threat to date. [7] The quantity of active virus within dental aerosols has not been confirmed and the amount of virus exposure which can result in infection is also currently unclear. At present, the World Health Organisation (WHO) recommends airborne precautions for AGPs in conjunction with undertaking risk assessments, frequent hand hygiene, respiratory etiquette, and environmental cleaning and disinfection. [1] Droplets/aerosol particles may transfer from the patient’s mouth to the breathing zone or body surface of the dental team, thereby contributing to the spread of infections. It is the ‘perfect storm’ of the routine production of these aerosols, combined with patients that might be asymptomatic carriers of SARS-CoV-2, plus the lack of good evidence regarding measures to mitigate the effects of dental aerosols that is at the root of the issues facing dental practitioners and dental schools. These impact dentists in terms of their clinical practise, use of PPE and an arbitrarily determined length of fallow time, leading to a reduction in the number of patients that can be treated. Following the suspension of routine dental treatment, advice was provided from dental councils around the world that AGPs should be stopped (avoid/defer) unless necessary e.g. for emergency procedures. [8] Various SOPs have since been published by a number of learned and professional organisations to guide dentists on safe working practice, however many of these acknowledge that there is only a limited evidence base.

Various methodologies for determining aerosolization in dental environments have been implemented, including the use air particle measurement, [9, 10] biological air sampling, [11, 12] culture of settle plates [6, 13, 14] and fluorescent markers. [9, 15, 16] Each of these methods offers insight into bioaerosol production, but each has its limitations. For instance, settle plates cannot account for the smallest particles that will not settle out of the air, air particle data cannot distinguish “clean” particles from the dental water unit line and those of biological origin, and the use of fluorescent dyes cannot offer information on viability of any potential biological component. Therefore, none of these methods alone can proffer robust findings as to the dispersal of active SARS-CoV-2.

Here we report a novel viral model for bioaerosol enumeration, utilising the bacteriophage Φ6 (Phi6) as a surrogate for SARS-CoV-2. Structurally the Φ6 virus particle is similar to SARS-CoV-2, in that it is a double-stranded RNA virus of approximately ∼80-100 nm in size, comprised of a lipid membrane envelope and spike proteins. [17] Recent literature has suggested Φ6 is an appropriate surrogate for infectious enveloped viruses, such as coronavirus. [18-20] This bacteriophage, from the Cystoviridae family of viruses, can be used in conjunction with the host bacterium *Pseudomonas syringae* to provide a valuable viral detection system. To our knowledge, this is the first study investigating the aerosolization of a bacteriophage, as an active biological marker and SARS-CoV-2 surrogate, in the dental environment.

The overall aim of this study is to use a multifaceted approach to the measurement of aerosol dispersal in a dental surgery setting, to determine the potential infection risk to the dental team from bioaerosol exposure during routine dental procedures and to investigate optimal mitigation strategies and the necessity of fallow time.

## Methodology

### Microorganism strains and culture conditions

*Pseudomonas syringae* (DSM 21482) and bacteriophage Φ6 (DSM 21518) were acquired from the German Collection of Microorganisms and Cell Cultures, Leibniz Institute, Germany. *P. syringae* was cultured on Tryptic Soy Agar (TSA; Sigma, UK) in 5% CO_2_ for 48 hours at 25°C (Forma Scientific, UK) or in Tryptic Soy Broth (TSB; Oxoid, UK) for 18 hours at 25°C, 150 rpm.

Bacteriophage Φ6 was propagated as previously reported, with modifications. [21, 22] Briefly, 300 µL of bacteriophage stock was added to 30 mL of TSB and 1 mL of *P. syringae* culture in exponential growth phase. The suspension was grown overnight at 25°C with orbital shaking at 150 rpm (MaxQ 4450, Thermo scientific, UK). The lysate was centrifuged at 3,250 rpm for 10 min at 4°C and the supernatant filtered (0.22 µm) to remove remaining bacterial cell debris. Φ6 bacteriophage suspensions were diluted 1:1 with SM buffer [0.1M NaCl, 8mM MgSO_4_. 7H2O, 50mM Tris-Cl, 0.01% w/v gelatin; pH7.5] and stored at 4°C. The titre was determined via the double-agar layer method as previously described. [22-24] Briefly, bacteriophage stock was serially diluted in SM buffer to 10^− 6^ plaque forming units (pfu). *P. syringae* was cultured overnight in TSB as previously and adjusted to an optical density at 600nm of 0.6 (OD_600_ 0.6) using a spectrophotometer (Jenway 6305, UK). 500 μL of each bacteriophage stock dilution and 200 μL of *P. syringae* culture were added to 6 mL of molten top-layer agar, consisting of 33g L^-1^ TSB and 6.6 g L^-1^ Agar (Sigma, UK), swirled and poured onto TSA plates and incubated (25°C, 18 hours). The Φ6 bacteriophage stock titre was approximately 10^8^ pfu mL^-1^.

### Experimental model design

Experiments were conducted at the Leeds Dental Institute (LDI), UK, in a clinical surgery with a design airflow of nine air changes per hour (ACH); measured by balometer (PH731 Capture hood, TSI AirFlow Instruments, USA) at 8.3 ACH by the investigators. A dental phantom head (Nissin Dental Products Inc., Japan) adapted to fit a dental chair, acted as a surrogate patient and tooth preparations were performed on hard thermosetting plastic teeth (Frasaco, Germany) in the UL2 and UL6 position. A flow of artificial saliva (mucin 1.8 g L^-1^, NaCl 0.29 g L^-1^, KH_2_PO_4_ 0.84 g L^-1^, ascorbic acid 0.00015 g L^-1^, urea 0.41 g L^-1^, arginine 0.65 g L^-1^, proteose peptone 3.3 g L^-1^, tryptose peptone 1.5 g L^-1^, L-cysteine hydrochloride 0.045 g L^-1^ (pH7.4)), containing Φ6 bacteriophage (∼10^8^ pfu mL^-1^) was introduced from three anatomical positions, 2x parotid, 1x sublingual (Figure 1B). These positions were used for endodontic access procedures on the upper first molar tooth. For the anterior crown preparation, the saliva port from the upper left first molar was moved to over the apex of the upper left lateral incisor. Saliva was flowed continuously into the oral cavity through 1 mm diameter tubing and a 120S/DM2 peristaltic pump (Watson Marlow, UK). The total salivary flow was 1.5 mL min^-1^, split equally across the three positions. Prior to commencing the procedures, the surfaces of the oral cavity were coated with artificial saliva containing the bacteriophage.

**Figure 1.**
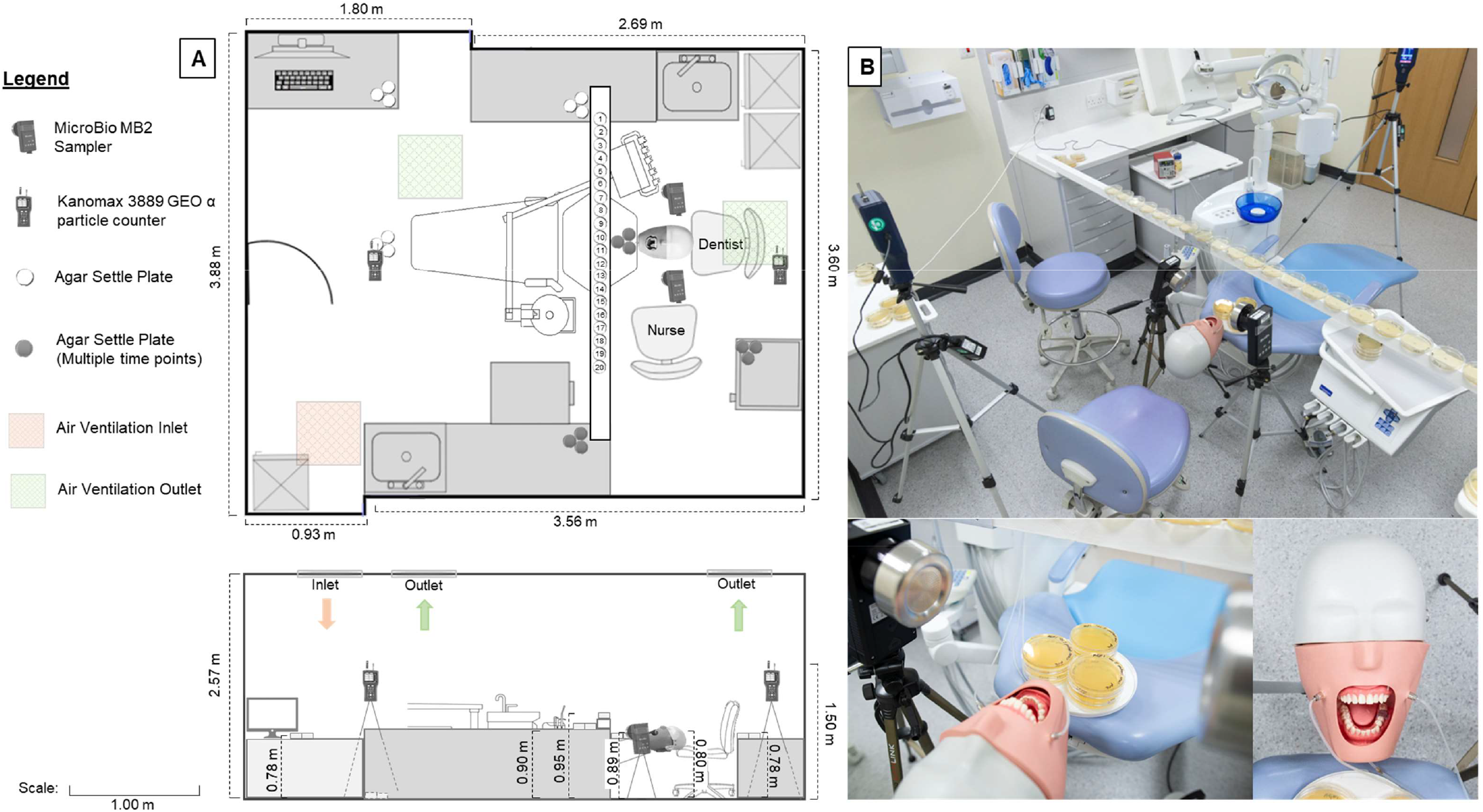
Experimental setup in the Leeds Dental Institute surgery. A – Schematic of experimental setup, top - above, bottom – side view. B – Top – Photographic overview of surgical clinic layout, Bottom – Close up of the phantom head with saliva tubing inputs and oral cavity configuration.

### Aerosol generating procedures

The AGPs investigated were root canal access of the upper left first molar and a full crown preparation on the upper left lateral incisor, with an assistant providing high-volume aspiration where necessary. A KaVo EXPERTtorque LUX E680L air turbine hand-piece (KaVo Dental GmbH, Germany) with an approximate cutting speed of 200,000 rpm or a NSK S-Max M95L electric speed-controlled hand-piece (Nakanishi Inc., Japan) at 60,000 rpm were used. The endodontic access was carried out with a pear-shaped fissure bur (Hidi-Once Diamond Bur 526 Med; Dentsply, USA). Several mitigation strategies were assessed including high-volume aspiration, rubber dam and aspiration and a specialist aerosol extraction device, Aspi Jet 25 with flute-shaped end piece (Cattani Air Technology, Italy), used as per manufacturer’s instructions, in the 6 o’clock position. Each experimental procedure was performed a minimum of three times and comprised of a 10-minute settle period post-setup, 20-minute AGP, followed by 20 minutes fallow time. The AGP consisted of four minutes of active hand-piece, followed by one minute of rest, repeated four times for a total AGP time of 20 minutes.

### Bacteriophage dispersal detection

Both passive and active sampling was undertaken to monitor the spread of bioaerosol during the AGPs. Settle plates and microbiological air sampling plates containing the bacteriophage host (*P. syringae*) were used to detect aerosol and droplets in the environment. Standard petri dish (90 mm diameter) settle plates consisting of TSA supplemented with 50 mg L^-1^ cycloheximide (Fisher Scientific, UK) were lawned with 200 µL of *P. syringae* culture (OD_600_ 0.6) and allowed to air dry. Triplicate settle plates were positioned in specific locations of the dental surgery, at either breathing zone, bench height or on the floor (Figure 1A). In three locations settle plates were exposed at specific time points of the procedures (during AGP and post-AGP fallow period). A series of 20 settle plates arranged across the surgery, proximal to the oral cavity (Figure 1), was used to collect bioaerosol and measure the distance that splatter (particles larger than 50µm) [5] and aerosolized droplets travelled.

Further *P. syringae*-lawned settle plates were used with two MicroBio MB2 air sampling devices (Cantium Scientific, UK), set 30 cm either side of the oral cavity (Figure 1). These devices draw air onto the *P. syringae*-lawned plates at 100 L min^-1^ and were set to sample 400 L of air during each of the four four-minute periods when the dental hand-piece was active. Two further, delayed air samples of 400 L were taken during the fallow period, between the six and ten-minute timepoints. Air sample counts were adjusted by a positive hole correction factor. [25] Procedures were carried out in series, to enable different mitigation strategies to be compared with each other and a no mitigation baseline. Fresh PPE was donned for each procedure to prevent cross contamination between experiments and only the dentist and investigator (also acting as dental nurse) were present during AGPs. Post-procedure, a third investigator (wearing PPE) sealed settle plates as a final anti-cross contamination measure.

### Air particle enumeration and size determination

Two Kanomax 3889 GEO α particle counters (Kanomax, Japan) were used to monitor size and quantity of particles in six size ranges simultaneously; diameters 0.3 μm, 0.5 µm, 1.0 μm, 3.0 µm, 5.0 μm and 10 μm. One particle counter was positioned directly behind the dentist, and another was stationed between the dental chair and the door (Figure 1), to monitor the generation of aerosols prior to, during and post-AGP. Counters were situated at a height of 150 cm, corresponding to the average adult breathing zone. Measurements (particles/m^3^) were recorded in one-minute repeating periods. Particle data was presented as baseline-standardised readings, where the counts for the final minute of the settle period were subtracted from the mean average of AGP levels.

### Statistical Analyses

Statistical analyses were performed using IBM SPSS Statistics 26. Direct comparisons of air turbine and electric hand-pieces and procedure location were performed using the Mann-Whitney U test. For bacteriophage data, Bonferroni corrections for multiple comparisons significance cut-off of *p=0*.*017*, whilst the cut-off for particle data was *p=0*.*008*.

## Results

### Bioaerosol dispersal during AGPs with a series of mitigation strategies

Bioaerosol was detected at all sampling points with an air turbine and no mitigation control (Figure 2). Each applied mitigation reduced levels of bioaerosol recovered from settle plates and air samples (Table 1), with the exception of the Aspi Jet 25 aerosol extraction device. The use of a rubber dam vastly reduced aerosolized bacteriophage and splatter. Across all mitigations for anterior crown preparations, the electric speed-controlled hand-piece generated significantly less bioaerosol than the air turbine; (*p<0*.*001*). For high-volume aspiration procedures, the electric speed-controlled hand-piece reduced 100.00% of settled aerosol (*p=0*.*037*) and 99.98% of bioaerosol recovered in air samples (*p=0*.*046*) compared with the air turbine. For no mitigation controls and rubber dam, settled bioaerosol counts were reduced with the electric speed-controlled hand-piece by 99.72% and 100.00%. Bacteriophage pfu detected through air sampling were significantly reduced by 99.49% and 100.00% for the same mitigation strategies (*p<0*.*001*).

**Table 1:** Mean Φ6 bacteriophage plaque forming units (pfu) collected with upper left lateral incisor procedures on settle and air sampling agar plates. Data is delineated by hand-piece and mitigation strategy and presented ± standard error of mean. Splatter zone radius defined within 41cm radius of mouth

|  | Total pfu (unless stated i.e. air samples) |  |  |  |  |  |  |
| --- | --- | --- | --- | --- | --- | --- | --- |
|  | Air Turbine |  |  |  | Electric Speed-Controlled |  |  |
|  | No mitigation (n=8) | High Volume Aspiration (n=3) | Dam (n=3) | Aerosol Extraction Device (n=3) | No mitigation (n=7) | High Volume Aspiration (n=3) | Dam (n=3) |
| Splatter Zone | 1152.75<br>($\pm 345.50$) | 1455.00<br>($\pm 190.05$) | 106.67<br>($\pm 35.53$) | 1255.33<br>($\pm 245.19$) | 54.43<br>($\pm 20.59$) | 64.33<br>($\pm 61.84$) | 5.33<br>($\pm 3.53$) |
| Settled aerosol | 207.00<br>($\pm 98.22$) | 90.33<br>($\pm 26.09$) | 0.33<br>($\pm 0.33$) | 86.33<br>($\pm 22.64$) | 0.57<br>($\pm 0.57$) | 0.00<br>($\pm 0.00$) | 0.00<br>($\pm 0.00$) |
| Average Air (pfu/m<sup>3</sup>) | 940.98<br>($\pm 56.29$) | 637.40<br>($\pm 142.79$) | 1.35<br>($\pm 0.27$) | 380.10<br>($\pm 68.74$) | 4.82<br>($\pm 1.29$) | 0.10<br>($\pm 0.04$) | 0.00<br>($\pm 0.00$) |
| Fallow settle | 24.75<br>($\pm 16.72$) | 21.00<br>($\pm 18.01$) | 0.00<br>($\pm 0.00$) | 2.00<br>($\pm 0.58$) | 0.57<br>($\pm 0.57$) | 0.00<br>($\pm 0.00$) | 0.00<br>($\pm 0.00$) |
| Average Fallow Air (pfu/m<sup>3</sup>) | 29.38<br>($\pm 2.80$) | 11.25<br>($\pm 0.29$) | 0.00<br>($\pm 0.00$) | 2.92<br>($\pm 0.17$) | 0.00<br>($\pm 0.00$) | 0.00<br>($\pm 0.00$) | 0.00<br>($\pm 0.00$) |

**Figure 2.**
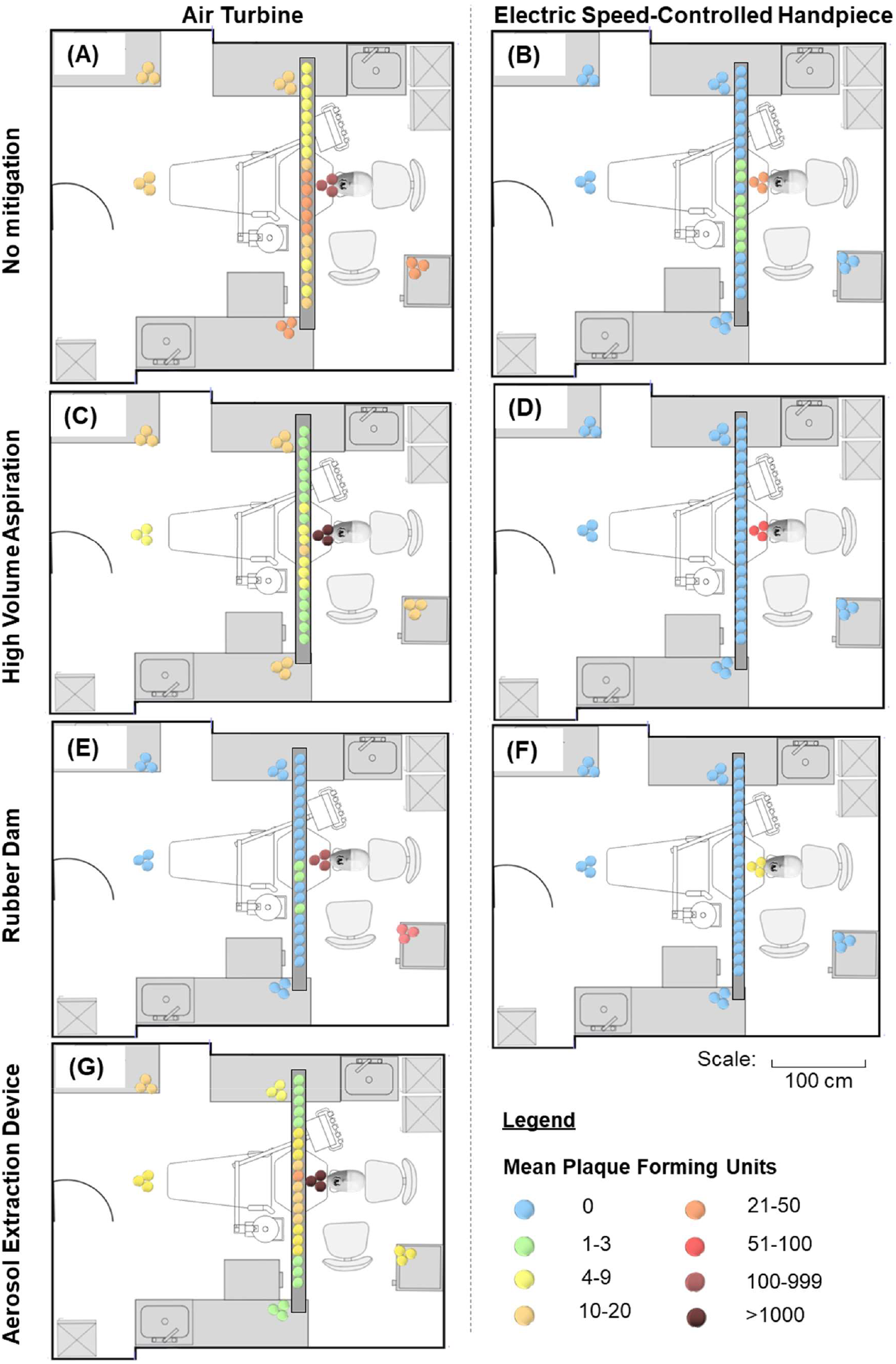
Bacteriophage dispersal heat maps by hand-piece and mitigation strategy. Data from anterior crown preparation procedures. A-B: no mitigation, C-D: high-volume aspiration, E-F: rubber dam, G: Aspi Jet 25.

Procedures employing the air turbine with high-volume aspiration demonstrated levels of bacteriophage of 21 pfu and 11.25 pfu/m^3^, for post-AGP settled bioaerosols and microbiological air samples respectively (Table 1). The use of an electric speed-controlled hand-piece reduced both to zero; (*p=0*.*037* and *p=0*.*037*, respectively). The use of rubber dam for either hand-piece also resulted in undetectable bioaerosol for the post-procedure, fallow period.

### Assessment of hand-piece and mitigation strategies through air particle analysis

Particles for all size ranges recorded in anterior procedures demonstrated reduced counts with the electric speed-controlled hand-piece versus air turbine, with the exception of 0.3 µm particles under rubber dam; (Figure 3, Appendix Table 1). Significant differences were observed between the hand-pieces with no mitigation for several particle size ranges; *p=0*.*017, p=0*.*005, p=0*.*001, p=0*.*008, p=0*.*021, p=0*.*059* for 0.3 µm, 0.5 µm, 1.0 µm, 3.0 µm, 5.0 µm, 10.0 µm, respectively.

**Figure 3.**
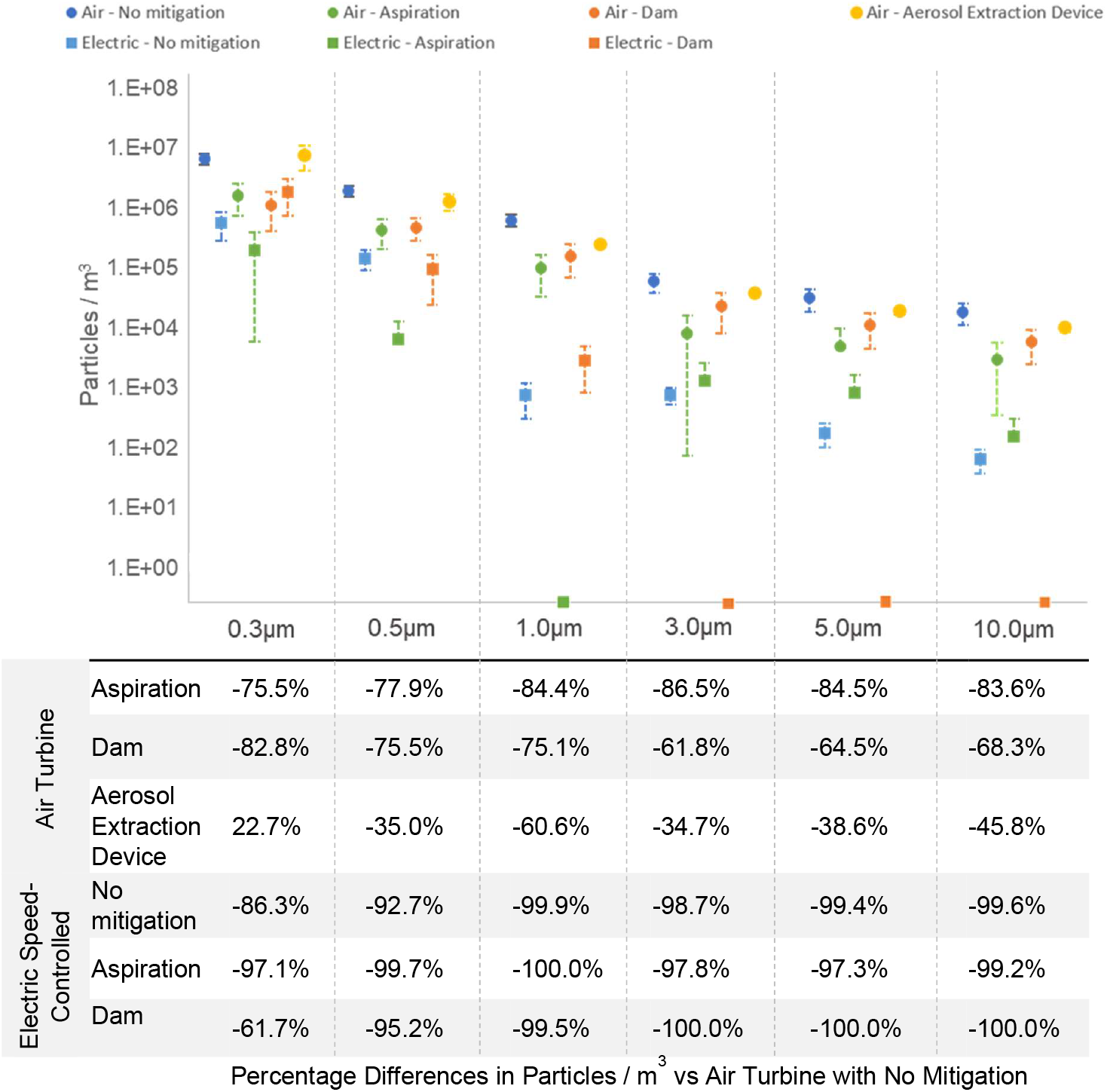
Top – Mean particles / m^3^ by size, produced during aerosol-generating procedures with air turbine and electric speed-controlled hand-pieces and a series of mitigation strategies. Data is standardised by the baseline control. Bottom - Percentage differences in particles / m^3^ produced during a series of procedures versus air turbine and no mitigation. Data from the behind dentist location and the anterior position.

The length of time, post-AGP for particle levels to return to pre-procedure baseline levels was highly variable, with 22.2% of procedures not reaching baseline figures within 25 minutes. There were no discernible differences between air turbine and electric hand-pieces for the time to return to baseline for 0.3 µm and 0.5 µm particles, 16.3 vs 18.2 and 15.7 vs 17.2 minutes, respectively. Differences were more apparent for 1.0 µm, 3.0 µm, 5.0 µm and 10.0 µm particles, demonstrating average times to reach to baseline of 16.7 vs 7.4, 14.1 vs 4.3, 12.6 vs 3.9 and 12.8 vs 4.5 minutes, respectively.

### Assessment of bioaerosol generation in anterior versus posterior tooth positions

Both settled and air bioaerosol measurements demonstrated decreases of >92% bacteriophage counts when comparing UL6 versus UL2 positions, with aspiration reducing settle by 99.6% and air sample readings by 100.0% (Table 2). Particle data was also vastly reduced in posterior endodontic experiments versus anterior procedures for all particle sizes and sampling positions; (Table 3, Appendix Table 2).

**Table 2:** Mean average Φ6 bacteriophage plaque forming units (pfu) collected during procedures at the UL6 position compared to data from the UL2 position. Data is delineated by mitigation strategy and presented ± standard error of mean. Splatter zone radius defined within 41cm radius of mouth. * both pfu counts were zero. p value based on Mann-Whitney U Test.

|  | No mitigation (n=4) |  |  | High-Volume Aspiration (n=3) |  |  | Dam (n=3) |  |  |
| --- | --- | --- | --- | --- | --- | --- | --- | --- | --- |
|  | UL6 Total pfu (unless stated) | Percentage Difference to UL2 (%) | UL6 vs UL2 (p value) | UL6 Total pfu (unless stated) | Percentage Difference to UL2 (%) | UL6 vs UL2 (p value) | UL6 Total pfu (unless stated) | Percentage Difference to UL2 (%) | UL6 vs UL2 (p value) |
| Splatter Zone | 68.00 (±51.88) | -94.1% | p<0.01 | 37.67 (±31.39) | -97.4% | p=0.10 | 253.67 (±215.38) | 137.8% | p=1.00 |
| Settled aerosol | 2.75 (±1.11) | -98.7% | p<0.01 | 0.33 (±0.33) | -99.6% | p=0.10 | 0.00 (±0.00) | -100.0% | p=0.70 |
| Air (pfu/m<sup>3</sup>) | 1.25 (±0.51) | -99.7% | p<0.01 | 0.00 (±0.00) | -100.0% | p=0.10 | 0.04 (±0.04) | -92.3% | p=0.40 |
| Fallow settle | 0.00 (±0.00) | -100.0% | p=0.02 | 0.00 (±0.00) | -100.0% | p=0.10 | 0.00 (±0.00) | 0* | p=1.00 |
| Fallow Air (pfu/m<sup>3</sup>) | 0.13 (±0.13) | -98.9% | p=0.01 | 0.00 (±0.00) | -100.0% | p=0.10 | 0.00 (±0.00) | 0* | p=1.00 |

**Table 3:** Percentage differences in particles / m^3^ produced during air turbine procedures at the UL6 position compared to data from the UL2 position. Data is delineated by mitigation strategy and sampling position. Dentist position is 115cm from the mouth, behind the practitioner. Door position is 185cm from the mouth past the foot of the chair. Splatter zone radius defined within 41cm radius of mouth.

|  |  |  | No mitigation | Aspiration | Dam |
| --- | --- | --- | --- | --- | --- |
| Percentage Difference in Particles / m<sup>3</sup><br>(UL6 vs UL2) | 0.3 µm | Dentist | -73% | -89% | -70% |
|  |  | Door | -81% | -15% | -77% |
|  | 0.5 µm | Dentist | -82% | -97% | -77% |
|  |  | Door | -86% | -37% | -84% |
|  | 1.0 µm | Dentist | -85% | -100% | -80% |
|  |  | Door | -89% | -76% | -83% |
|  | 3.0 µm | Dentist | -89% | -100% | -84% |
|  |  | Door | -87% | -94% | -84% |
|  | 5.0 µm | Dentist | -94% | -100% | -96% |
|  |  | Door | -89% | -60% | -86% |
|  | 10.0 µm | Dentist | -96% | -84% | -85% |
|  |  | Door | -95% | -55% | -88% |

## Discussion

To model the spread of bioaerosols during AGPs we performed our experiments with a high salivary bacteriophage concentration of ∼10^8^ pfu, close to maximum reported physiological levels of SARS-CoV-2 in human saliva. [26, 27] For the endodontic access on the upper left molar tooth, saliva was introduced into the mouth at sites representative of the parotid and sublingual saliva glands. For the upper left lateral incisor crown preparation, saliva was introduced above the apex of the tooth, representing a worse-case scenario. Different mitigation strategies using both air turbine and electric hand-piece were used for each dental procedure. There was a clear distinction between the amount of aerosolized saliva dispersed around the dental surgery using the traditional air turbine and an electric speed-controlled hand-piece. Bioaerosol levels were clearly diminished when using the electric hand-piece over the air turbine. Whilst the electric hand-piece was used at 60,000 rpm, compared to higher speeds with the air turbine, the absence of compressed air appears to be key to a reduced bioaerosol production.

Air sampling was also implemented with the bacteriophage methodology to ensure the quantification of viable viral particles in the air, potentially too small to settle onto the clinic surfaces. [28] With high-volume aspiration, the differences displayed between air and electric hand-piece AGPs were large, 637.4 pfu/m^3^ versus 0.1, respectively. The latter represented a solitary bacteriophage unit detected in a single experimental replicate. For all mitigations, the settle plates closest to the mouth indicated the highest quantities of bacteriophage (Figure 2). This was due to splatter rather than bioaerosol and can be consider a lower transmission risk than aerosolized virus. The data in this study were collected in an environment with mechanical ventilation. Whilst many “high street” practices around the world do not have these facilities, evidence indicates that aerosol accumulation is greater in practices with poor ventilation. [29]

These differences are clearly supported by the particle measurement data, which indicated lower levels of all particles sizes for the majority of mitigations; (Figure 3). Furthermore, by recording particle measurements from two locations in the surgery, we saw that aerosols were not localised and displayed similar, although slightly delayed trends, towards the extremities of the clinic. This further highlights the necessity for good mitigation protocols.

The evidence presented here also corroborates data reported by others, suggesting that the use of a rubber dam significantly reduces microbial aerosolization. [30, 31] Whilst these studies report bacterial air contamination rates, our study documents the reduction of viral aerosolization, which may be expected to behave differently due to their smaller size. Conversely, the Aspi Jet 25, specialist aerosol extraction device was no better than high-volume aspiration alone. Although it reduced the biological particles in the air, it seemed to increase air turbulence without substantially removing the majority of bioaerosol.

To improve the accuracy of the phantom head unit over previous published models, [9, 15, 32] we added an anatomical tongue model and a high-level physiological salivary flow of 1.5 mL min^-1^, spread across three positions. Whilst an improvement on models reported in the literature, it is not without its limitations. The model uses artificial teeth, which lack the anatomical intricacies of human teeth. However, particle analysis by Shahdad et al reported little difference between plastic and real teeth. [32] Our model does not imitate the effects of a patient breathing, or other patient behaviours, such as talking or coughing which would likely contribute considerably to bioaerosol production. Nonetheless, by demonstrating reductions in active biological marker dispersal, we can make robust indications as to the value of dental mitigation approaches. Whilst other approaches to bioaerosol measurements, such as the Gesundheit II, [33] are available, this device does not permit the assessment of dispersal throughout a room.

The requirement for, and length of, a period of fallow time is far from clear, with estimates ranging from two to 180 minutes. [32, 34, 35] However, this is often based on particle data alone. When based on particle count data, Ehtezazi et al suggested that with no mitigation the average time to return to baseline levels was between 28 and 34 minutes. [35] However, this study had no salivary element, which potentially contributes significantly to particle levels. In this study, we determined a wide variation in the time it took for particles levels of all sizes to return to pre-AGP levels, although the larger particles required less time with the electric hand-piece procedures. Shahdad et al reported similar variability, also suggesting that fallow time estimates were longer for procedures where the hand-piece was used in five minute bursts, [32] comparable to the protocol used in this experiment. Considering other methods to assess fallow time, a recent study from Allison et al indicated that <0.1% of aerosolized fluorescein dye was detectable after 30 minutes fallow time. [9] Whilst spectrofluorometric analysis such as this can provide valuable information, it does not inform about the viability of the particles transferred. The bacteriophage data in this study demonstrated that with air turbine and high-volume aspiration, substantial amounts of both settle and aerosolized bacteriophage were detectable between the six and ten minute fallow period; (Table 1). However, use of the electric speed-controlled hand-piece eliminated any bioaerosol within six minutes of the procedure completion. Therefore, this evidence strongly suggests there is no need for a prolonged fallow period with this hand-piece. Where an electric hand-piece is not available, the use of a rubber dam was equally effective in reducing air contamination shortly after conclusion of an AGP. Assessing both the particle and bacteriophage fallow data together, it becomes clear that particle data alone cannot provide sufficient information to determine risk of airborne viral particles. Here we see instances of baseline levels not being achieved post-AGP, but no detectable active bacteriophage in the air.

We assessed the differences between anterior and posterior AGP positions. Both the bacteriophage and particle data highlighted the importance of procedural position on risk. When using an air turbine and high-volume aspiration, no bacteriophage particles were detected in the air during the posterior procedures with a >84% reduction in all particle sizes observed from behind the dentist. Together, these data support the interpretation that endodontic procedures in the posterior of the mouth impart a lower risk of viral contamination and dispersal into the environment, with bioaerosols most likely trapped inside the oral cavity. Conversely, dental procedures in the anterior region pose the greatest risk.

Through the combined use of novel and established methodologies, the data described here presents a clear picture of how risk of SARS-CoV-2 and other similar biological hazards can be greatly attenuated by the use of electric speed-controlled hand-pieces. Whilst detection of a single viral unit may not translate to an infective viral load, the reduction in levels with these mitigating approaches is clear to see. This study further suggests that with these hand-pieces and high-volume aspiration or the use of rubber dam, a prolonged fallow period is not necessary in the clinical setting used. Equipping our dental surgeries with these tools will be crucial to protecting the health, safety and future of dental teams and services. Finally, the data presented here suggests that particle count data alone cannot provide accurate information as to the dispersal and settlement of bioaerosols, with bacteriophage markers offering a greater insight into infection risk.

## Data Availability

All data is presented in the article. No patient data was used.

## Author Contributions

JV contributed to experimental design, data collection, analysis, manuscript writing and manuscript revision.

EB contributed to experimental design, data collection and manuscript revision.

TD contributed to experimental design, data collection and manuscript revision.

DD contributed to study conception, experimental design, analysis and manuscript revision.

LF contributed to study conception, experimental design, data collection, analysis and manuscript revision.

DW contributed to study conception, experimental design, data collection, analysis and manuscript revision.

BN contributed to study conception, experimental design, data collection, analysis and manuscript revision.

## Acknowledgements

We would like to thank the Leeds Dental Institute for accommodating the requirements for clinical space to perform these experiments, Dr Jing Kang for advice on statistical analyses and Tim Zoltie for photography.

