## Appendix Table for "Dental mitigation strategies to reduce aerosolization of SARS-CoV-2"

| UL2 – Dentist Position | |  | 0.3 µm | 0.5 µm | 1.0 µm | 3.0 µm | 5.0 µm | 10.0 µm |
| --- | --- | --- | --- | --- | --- | --- | --- | --- |
| Air Turbine | No mitigation | Mean | 6.54E+06 | 1.97E+06 | 6.14E+05 | 6.04E+04 | 3.06E+04 | 1.66E+04 |
|  |  | ± STD Error | 1.31E+06 | 4.03E+05 | 1.31E+05 | 2.13E+04 | 1.23E+04 | 6.71E+03 |
|  | High-Volume Aspiration | Mean | 1.66E+06 | 4.36E+05 | 9.59E+04 | 8.14E+03 | 4.74E+03 | 2.72E+03 |
|  |  | ± STD Error | 2.88E+05 | 5.33E+04 | 4.36E+02 | 2.33E+02 | 7.57E+01 | 2.52E+01 |
|  | Dam | Mean | 1.12E+06 | 4.84E+05 | 1.53E+05 | 2.31E+04 | 1.08E+04 | 5.26E+03 |
|  |  | ± STD Error | 9.15E+05 | 2.22E+05 | 6.32E+04 | 8.07E+03 | 4.74E+03 | 2.40E+03 |
|  | Aerosol Extraction Device | Mean | 7.61E+06 | 1.28E+06 | 2.42E+05 | 3.94E+04 | 1.88E+04 | 9.01E+03 |
|  |  | ± STD Error | 1.95E+05 | 6.59E+03 | 0.00E+00 | 1.34E+03 | 8.17E+02 | 1.40E+02 |
| Electric Rotor | No mitigation | Mean | 5.73E+05 | 1.44E+05 | 7.36E+02 | 7.73E+02 | 1.77E+02 | 5.88E+01 |
|  |  | ± STD Error | 7.04E+05 | 1.95E+05 | 8.55E+04 | 1.49E+04 | 6.45E+03 | 3.05E+03 |
|  | High-Volume Aspiration | Mean | 2.01E+05 | 6.59E+03 | 0.00E+00 | 1.34E+03 | 8.17E+02 | 1.40E+02 |
|  |  | ± STD Error | 1.14E+06 | 7.09E+04 | 1.99E+03 | 0.00E+00 | 0.00E+00 | 0.00E+00 |
|  | Dam | Mean | 1.90E+06 | 9.55E+04 | 2.80E+03 | 0.00E+00 | 0.00E+00 | 0.00E+00 |
|  |  | ± STD Error | 3.32E+06 | 3.88E+05 | 2.44E+04 | 3.83E+03 | 2.38E+03 | 1.48E+03 |
| UL2 – Door Position | |  | 0.3 µm | 0.5 µm | 1.0 µm | 3.0 µm | 5.0 µm | 10.0 µm |
| Air Turbine | No mitigation | Mean | 6.63E+06 | 1.96E+06 | 5.86E+05 | 4.59E+04 | 2.30E+04 | 1.12E+04 |
|  |  | ± STD Error | 1.05E+06 | 2.57E+05 | 7.58E+04 | 1.01E+04 | 5.59E+03 | 2.76E+03 |
|  | High-Volume Aspiration | Mean | 1.70E+06 | 4.14E+05 | 8.37E+04 | 5.66E+03 | 2.17E+03 | 8.48E+02 |
|  |  | ± STD Error | 8.56E+05 | 2.39E+05 | 5.61E+04 | 5.00E+03 | 2.17E+03 | 8.48E+02 |
|  | Dam | Mean | 1.16E+06 | 4.33E+05 | 1.29E+05 | 1.45E+04 | 6.91E+03 | 2.24E+03 |
|  |  | ± STD Error | 6.01E+05 | 9.97E+04 | 5.37E+04 | 8.90E+03 | 4.03E+03 | 1.51E+03 |
|  | Aerosol Extraction Device | Mean | 7.79E+06 | 1.37E+06 | 2.27E+05 | 3.66E+04 | 1.82E+04 | 9.09E+03 |
|  |  | ± STD Error | 3.33E+06 | 5.67E+05 | 3.25E+04 | 4.56E+03 | 2.61E+03 | 1.43E+03 |
| Electric Rotor | No mitigation | Mean | 3.49E+05 | 1.63E+05 | 3.96E+03 | 1.10E+03 | 6.50E+02 | 4.26E+02 |
|  |  | ± STD Error | 1.45E+05 | 1.33E+05 | 3.96E+03 | 9.88E+02 | 5.83E+02 | 3.94E+02 |
|  | High-Volume Aspiration | Mean | 0.00E+00 | 0.00E+00 | 0.00E+00 | 3.13E+02 | 3.84E+02 | 3.55E+02 |
|  |  | ± STD Error | 0.00E+00 | 0.00E+00 | 0.00E+00 | 3.13E+02 | 3.84E+02 | 3.55E+02 |
|  | Dam | Mean | 5.80E+05 | 5.58E+04 | 5.64E+02 | 0.00E+00 | 0.00E+00 | 0.00E+00 |
|  |  | ± STD Error | 3.55E+05 | 5.58E+04 | 5.64E+02 | 0.00E+00 | 0.00E+00 | 0.00E+00 |

Appendix Table 1: Air particle counts standardized against the baseline data for “Dentist” and “Door” positions for different hand-pieces and mitigation strategies during upper left 2 (UL2) procedures. STD – Standard error of the mean.

| UL6 –  Dentist & Door | |  | 0.3 µm | 0.5 µm | 1.0 µm | 3.0 µm | 5.0 µm | 10.0 µm |
| --- | --- | --- | --- | --- | --- | --- | --- | --- |
| UL6 - Air - Dentist | No mitigation | Mean | 1.78E+06 | 3.49E+05 | 9.14E+04 | 6.86E+03 | 1.93E+03 | 5.96E+02 |
|  |  | ± STD Error | 3.71E+05 | 4.34E+04 | 2.18E+04 | 1.73E+03 | 9.11E+02 | 5.96E+02 |
|  | High-Volume Aspiration | Mean | 1.89E+05 | 1.43E+04 | 0.00E+00 | 0.00E+00 | 0.00E+00 | 4.41E+02 |
|  |  | ± STD Error | 1.10E+05 | 1.30E+04 | 0.00E+00 | 0.00E+00 | 0.00E+00 | 4.41E+02 |
|  | Dam | Mean | 3.38E+05 | 1.10E+05 | 3.10E+04 | 3.58E+03 | 4.05E+02 | 7.73E+02 |
|  |  | ± STD Error | 1.73E+05 | 1.04E+05 | 3.10E+04 | 2.68E+03 | 2.10E+02 | 3.91E+02 |
| UL6 - Air - Door | No mitigation | Mean | 1.25E+06 | 2.65E+05 | 6.59E+04 | 5.89E+03 | 2.47E+03 | 5.79E+02 |
|  |  | ± STD Error | 6.13E+05 | 1.08E+05 | 2.52E+04 | 2.11E+03 | 1.14E+03 | 2.01E+02 |
|  | High-Volume Aspiration | Mean | 1.44E+06 | 2.61E+05 | 2.03E+04 | 3.60E+02 | 8.68E+02 | 3.82E+02 |
|  |  | ± STD Error | 1.21E+06 | 2.26E+05 | 1.70E+04 | 3.60E+02 | 8.68E+02 | 3.82E+02 |
|  | Dam | Mean | 2.63E+05 | 6.88E+04 | 2.24E+04 | 2.27E+03 | 9.64E+02 | 2.80E+02 |
|  |  | ± STD Error | 1.46E+05 | 6.88E+04 | 2.24E+04 | 1.84E+03 | 4.94E+02 | 2.80E+02 |

Appendix Table 2: Air particle counts standardized against the baseline data for “Dentist” and “Door” positions for different hand-pieces and mitigation strategies during upper left 6 (UL6) procedures. STD – Standard error of the mean.
